# Effects of low versus high inspired oxygen fraction on myocardial injury after transcatheter aortic valve implantation: A randomized clinical trial

**DOI:** 10.1101/2023.01.19.23284775

**Authors:** Youn Joung Cho, Cheun Hyeon, Karam Nam, Seohee Lee, Jae-Woo Ju, Jeehoon Kang, Jung-Kyu Han, Hyo-Soo Kim, Yunseok Jeon

## Abstract

**Background:** Oxygen therapy is used in various clinical situation, but its clinical outcomes are inconsistent. The relationship between the fraction of inspired oxygen (F_I_O_2_) during transcatheter aortic valve implantation (TAVI) and clinical outcomes has not been well studied. We investigated the association of F_I_O_2_ (low vs. high) and myocardial injury in patients undergoing TAVI.

**Methods:** Adults undergoing transfemoral TAVI under general anesthesia were randomly assigned to receive F_I_O_2_ 0.3 or 0.8 during procedure. The primary outcome was the area under the curve (AUC) for high-sensitivity cardiac troponin I (hs-cTnI) during the first 72 h following TAVI. Secondary outcomes included the AUC for postprocedural creatine kinase-myocardial band (CK-MB), acute kidney injury and recovery, conduction abnormalities, pacemaker implantation, stroke, myocardial infarction, and in-hospital mortality.

**Results:** Between October 2017 and April 2022, 72 patients were randomized and 62 were included in the final analysis (n=31 per group). The median (IQR) AUC for hs-cTnI in the first 72 h was 42.66 (24.82–65.44) and 71.96 (35.38–116.34) h·ng/mL in the F_I_O_2_ 0.3 and 0.8 groups, respectively (p=0.066). The AUC for CK-MB in the first 72 h was 257.6 (155.6–322.0) and 342.2 (195.4–485.2) h·ng/mL in the F_I_O_2_ 0.3 and 0.8 groups, respectively (p=0.132). Acute kidney recovery, defined as an increase in the estimated glomerular filtration rate ≥ 25% of baseline in 48 h, was more common in the F_I_O_2_ 0.3 group (65% vs. 39%, p=0.042). Other clinical outcomes were comparable between the groups.

**Conclusions:** The F_I_O_2_ level did not have a significant effect on periprocedural myocardial injury following TAVI. However, considering the marginal results, a benefit of low F_I_O_2_ during TAVI could not be ruled out.

## Introduction

Although a fraction of inspired oxygen (F_I_O_2_) higher than that of ambient air is generally used during general anesthesia, there is continuing debate about the optimal F_I_O_2_. High oxygen tension is beneficial for reducing surgical site infection and in 2016 World Health Organization recommended that adults receive F_I_O_2_ 0.8 during mechanical ventilation under general anesthesia [1]. However, a more recent systematic review found no difference in the surgical site infection rate according to the intraoperative F_I_O_2_ amount, and suggested a negative effect of high F_I_O_2_ on long-term outcomes [2]. Other investigators did not find any difference in the degree of myocardial injury between perioperative F_I_O_2_ 0.3 and 0.8, and suggested that F_I_O_2_ 0.8 is safe for major non-cardiac surgery [3].

High oxygen tension may cause oxidative stress, coronary vasoconstriction, and altered microvascular perfusion, resulting in adverse systemic effects including myocardial injury [4]. In a meta-analysis of acute myocardial infarction (MI), there was no evidence to support the routine use of oxygen treatment and the authors could not rule out a harmful effect of unnecessary oxygen therapy [5]. Constant and brief intermittent hyperoxia both induced inflammatory responses and cytotoxicity in cardiomyocytes from adult humans [6]. Myocardial injury is common following transcatheter aortic valve implantation (TAVI) [7]. Moreover, abnormally increased cardiac biomarkers were associated with poor outcomes including periprocedural kidney injury and 30-day and 1-year mortality following TAVI [8].

The relationship between the F_I_O_2_ level and myocardial injury has not been well studied in patients undergoing TAVI. Therefore, this study investigated whether high (0.8) and low (0.3) F_I_O_2_ during transfemoral TAVI under general anesthesia have different effects on post-procedural myocardial injury, as indexed by serum cardiac troponin in the early post-TAVI period.

## Methods

### Ethics approval

This randomized controlled trial was approved by the Institutional Review Board of Seoul National University Hospital (#1707-109-871, on September 11, 2017) and registered at clinicaltrials.gov (NCT03291210, on September 25, 2017) before patient enrollment. The study was conducted according to the Good Clinical Practice guidelines and Declaration of Helsinki. Written informed consent was obtained from all participants, who could withdraw at any time.

### Study population and randomization

Adults (aged 20–99 years) with aortic stenosis (AS), undergoing elective TAVI under general anesthesia via the transfemoral approach in a single tertiary academic center (Seoul National University Hospital, South Korea), were eligible for the study. Eligibility for TAVI was based on the consensus of a local multidisciplinary heart team, including clinical cardiologists, cardiac interventionists, cardiac surgeons, radiologists, and anesthesiologists. The predicted operative mortality risk was calculated using the Society of Thoracic Surgeons Predicted Risk of Mortality (STS-PROM) score, European System for Cardiac Operative Risk Evaluation (EuroSCORE) II, and logistic EuroSCORE. The heart team determined the anesthetic method (general anesthesia or conscious sedation) based on the patients’ comorbidities, preference, and ability to maintain a supine position without profound dyspnea or restlessness during the procedure. The exclusion criteria were a non-transfemoral approach, pre-procedural arterial partial pressure of oxygen (PaO_2_) <65 mmHg or receiving oxygen treatment, severe pre-procedural renal dysfunction (defined as an estimated glomerular filtration rate [eGFR] <30 mL/min/1.73 m^2^), chronic pulmonary obstructive disease or symptomatic asthma, tuberculosis-destroyed lung, history of lung cancer, acute coronary syndrome within the past 6 months, documented pre-procedural cardiac troponin I (cTnI) or creatine kinase-myocardial band (CK-MB) elevation, stroke or transient ischemic attack within 6 months, pregnancy, and refusal to participate.

After enrollment and the informed consent process, the patients were randomized to receive F_I_O_2_ 0.3 or 0.8 during TAVI (1:1 allocation ratio) (Fig 1). Block randomization (blocks of four or six) was conducted using a computer-generated program by an independent research nurse on the morning of the intervention. The group assignments were concealed in an opaque envelop, and all investigators, patients, interventionists, and data analyzers were blinded to the group allocations.

**Fig 1.**
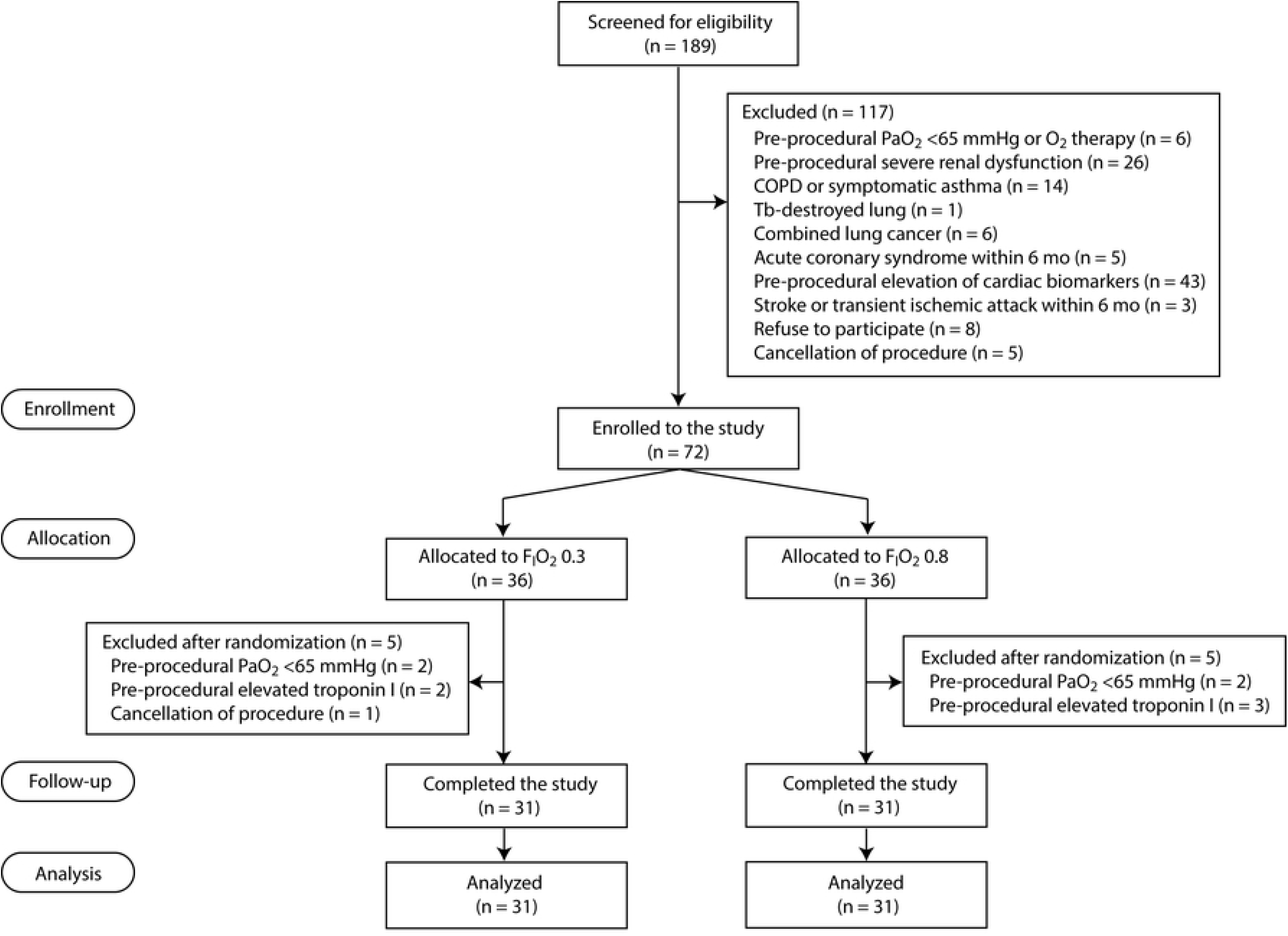
CONSORT diagram. CK-MB, creatine kinase-myocardial band; COPD, chronic obstructive pulmonary disease; F_I_O_2_, fraction of inspired oxygen; PaO_2_, arterial partial pressure of oxygen.

### Study protocol

The routine monitoring techniques of our institution for patients under general anesthesia were applied, except F_I_O_2_ management. Without premedication, 12-lead electrocardiogram, pulse oxygen saturation (SpO_2_), invasive and noninvasive arterial blood pressure, cerebral oxygen saturation (ScrbO_2_), and bispectral index monitoring were performed. Left and right ScrbO_2_ were measured using near-infrared spectroscopy (Somanetics INVOS oximeter; Covidien, Mansfield, MA, USA). Transesophageal or transthoracic echocardiography was performed to evaluate the valve position and presence of paravalvular regurgitation, as required by the interventionists.

Before inducing anesthesia, all participants were preoxygenated using an anesthesia machine (Primus; Drägerwerk, Lubeck, Germany) with F_I_O_2_ 0.3 or 0.8 according to the group allocation. After stabilization, general anesthesia was induced by a target-controlled infusion of propofol (effect-site concentration [Ce], 2.5–4.0 μg/mL) and remifentanil (Ce, 1.0–4.0 ng/mL) using a commercial infusion pump (Orchestra, Fresenius Vial, Brézins, France). Neuromuscular blockade was established by administering rocuronium (0.6 mg/kg). Then the trachea was intubated and the lungs were ventilated in volume-controlled mode with a tidal volume of 0.6–0.8 mL/kg and ventilatory rate of 9–12 /min. The alveolar recruitment maneuver was performed at 25 cmH_2_O for 10 s after tracheal intubation, and a positive end-expiratory pressure of 5 cmH_2_O was applied in all patients. According to the group assignment, F_I_O_2_ was maintained at 0.3 or 0.8 until the end of the TAVI procedure, unless the SpO_2_ was <93%. If desaturation occurred, F_I_O_2_ was increased by 0.05–0.1, and an additional alveolar recruitment maneuver was performed as needed to maintain SpO_2_ ≥93% by the attending anesthesiologists. On completing the procedure, 100% O_2_ was provided to all patients during anesthesia emergence. Patients were extubated in the intervention room, monitored in the cardiovascular care unit for 1–2 days, and then transferred to a general ward. Patients were discharged 5–7 days post-TAVI if they had no procedure-related complications.

The TAVI was conducted in accordance with the standard procedures in our institution. Using a transfemoral approach, a balloon-expandable Sapien III valve (Edwards Lifesciences, Irvine, CA, USA), self-expandable Evolut Pro or R valve (Medtronic, Minneapolis, MN, USA), or Lotus valve (Boston Scientific, Natick, MA, USA) was implanted at the diseased aortic valve. The valve was chosen by the heart team based on the size and structure of the native valve and sinus, heights of the coronary artery openings, and considerations regarding future coronary access, the risk of conduction disturbances, and annular calcification. The iliofemoral arteries were accessed under fluoroscopic guidance and closed percutaneously using Perclose ProGlide vascular suture-mediated closure devices (Abbott Vascular Devices, Redwood City, CA, USA). Before the procedure, the patients were given loading doses of dual antiplatelet agents: acetylsalicylic acid and clopidogrel (both 300 mg). During the procedure, the patients were heparinized with unfractionated heparin to achieve an activated clotting time >250 s. At completion of the valve implantation, the effects of heparin were reversed by protamine infusion.

During the procedure, arterial blood gas analysis (ABGA) was performed at four time points: baseline (before anesthesia induction, T1), after inducing general anesthesia (T2), after valve implantation (T3), and at the end of the procedure (T4). ABGA was performed using a GEM® Premier 3000 device (Model 5700; Instrumentation Laboratory, Lexington, MA, USA).

Two serum cardiac biomarkers of myocardial injury, high-sensitivity cTnI (hs-cTnI) and CK-MB, were measured at baseline (before the procedure) and 1, 4, 8, 24, 48, and 72 h after TAVI. hs-cTnI was measured using an Abbott Architect Plus Analyzer (i2000SR; Flex, San Jose, CA, USA), which has a limit of detection of 0.0011 μg/L and limit of blank of 0.0007–0.0013 μg/L. An hs-cTnI concentration ≥99^th^ percentile in the normal population (0.028 μg/L) was deemed abnormal. Serum creatinine concentrations were calibrated using isotope dilution mass spectrometry (IDMS). The eGFR was calculated using the modified diet in renal disease (MDRD) equation [9].

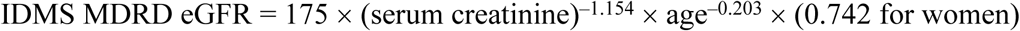

Postprocedural acute kidney injury (AKI) was determined based on the serum creatinine level and urine output according to the Kidney Disease: Improving Global Outcomes Clinical Practice Guidelines criteria for AKI [10]. AKI was defined as an increase in serum creatinine ≥1.5 times the baseline level or by ≥0.3 mg/dL (≥26.5 μmol/L) [AKI_creatinine_], or a urine output <0.5 mL/kg/h for ≥6 h within 7 days [AKI_urine output_]. AKI occurring >7 days after the procedure was excluded because it might have been unrelated to the procedure. Acute kidney recovery (AKR) was defined as an increase in eGFR of ≥25% relative to baseline at 48 h post-TAVI [11].

The postprocedural development of new conduction abnormalities and incidence of permanent pacemaker insertion was assessed. Stroke was defined as an acute episode of a focal or global neurological deficit as a result of hemorrhage or infarction, based on the Valve Academic Research Consortium-2 (VARC-2) definition [12]. Periprocedural MI was defined based on a combination of new ischemic symptoms or signs and elevated cardiac biomarkers within 72 h following TAVI, according to the VARC-2 definition [12].

### Study endpoints and sample size calculation

The primary study outcome was periprocedural myocardial injury, as reflected by the geometric area under the curve (AUC) for periprocedural serum hs-cTnI in the first 72 h post-TAVI, calculated according to the trapezoidal rule. Secondary outcomes were the AUC for serum CK-MB in the first 72 h post-TAVI, and the peak serum hs-cTnI and CK-MB levels in the same period. Post-procedural clinical outcomes were also evaluated, including AKI, AKR, new conduction abnormalities, permanent pacemaker insertion, stroke, MI, and in-hospital cardiovascular mortality.

To calculate the sample size, we conducted a pilot study of 10 patients undergoing transfemoral TAVI under general anesthesia. The AUC for serum hs-cTnI in the first 72 h after TAVI was 40.24 ± 28.16 ng/mL. Assuming that a 50% difference in hs-cTnI levels between the two treatment groups in the first 72 h is clinically relevant, 32 patients were required for each group at 80% power and an alpha error of 5%. Considering a 10% dropout rate, we recruited 36 patients to each group (a total of 72 patients).

### Statistical analysis

Data are presented as the mean ± SD, median (interquartile range, IQR), or number (%) after normality was tested using the Kolmogorov–Smirnov test. The primary endpoint, i.e., the AUC for serum hs-cTnI in the first 72 h after TAVI, was analyzed using the Mann–Whitney *U* test according to the data distribution. Other continuous variables were analyzed using the independent *t*-test or Mann–Whitney *U* test after performing a normality test. Categorical variables were analyzed using Pearson’s chi square test or Fisher’s exact test. For repeated measures, a linear mixed model with Bonferroni correction was used to compare the groups. In the mixed model, group, measurement time, and their interaction were fixed effects, while subject was a random effect. Plots of residuals versus fitted values were checked in terms of whether the error terms (residuals) had a mean of zero and constant variance. The normality assumption for repeated measures was assessed using histograms and quantile–quantile plots (for residuals). Multivariable logistic regression analysis was performed to identify independent risks for F_I_O_2_ and postprocedural hs-cTnI. Odds ratios (ORs) were adjusted for age, sex, STS-PROM score, C-reactive protein, left ventricular (LV) ejection fraction (EF), procedural time, and new conduction abnormality, and 95% confidence intervals (CIs) were presented. The analysis was done in an intention-to-treat manner. All analyses were performed using IBM SPSS Statistics (ver. 21.0; IBM Corp., Armonk, NY, USA) or R software (ver. 3.5.1; R Development Core Team, Vienna, Austria). A P value <0.05 was considered statistically significant.

## Results

Patients were screened for eligibility between October 18, 2017 and April 6, 2022. Of 189 patients, 117 were excluded based on the exclusion criteria (Fig 1). After 72 patients were randomized to the F_I_O_2_ 0.3 or 0.8 groups (n=36 each), 10 patients were excluded due to pre-procedural PaO_2_ <65 mmHg (n=4), elevated pre-procedural cTnI (n=5), or procedure cancellation (n=1). We noted violations of the exclusion criteria (pre-procedural PaO_2_ <65 mmHg or elevated cardiac biomarkers) in nine patients and excluded them from the analysis. Thus, a total of 62 patients (31 per group) were included in the final analysis. We performed additional sub-analysis including five patients (n=2 in the F_I_O_2_ 0.3 group and n=3 in the F_I_O_2_ 0.8 group) who had elevated cardiac biomarkers after randomization.

Tables 1 and 2 present the baseline characteristics of the included patients and procedural variables. Baseline characteristics were well balanced between the groups. The median (IQR) age of the included patients was 79 (77–83) years. The median (IQR) procedural duration was 80 (70–95) min.

**Table 1.**
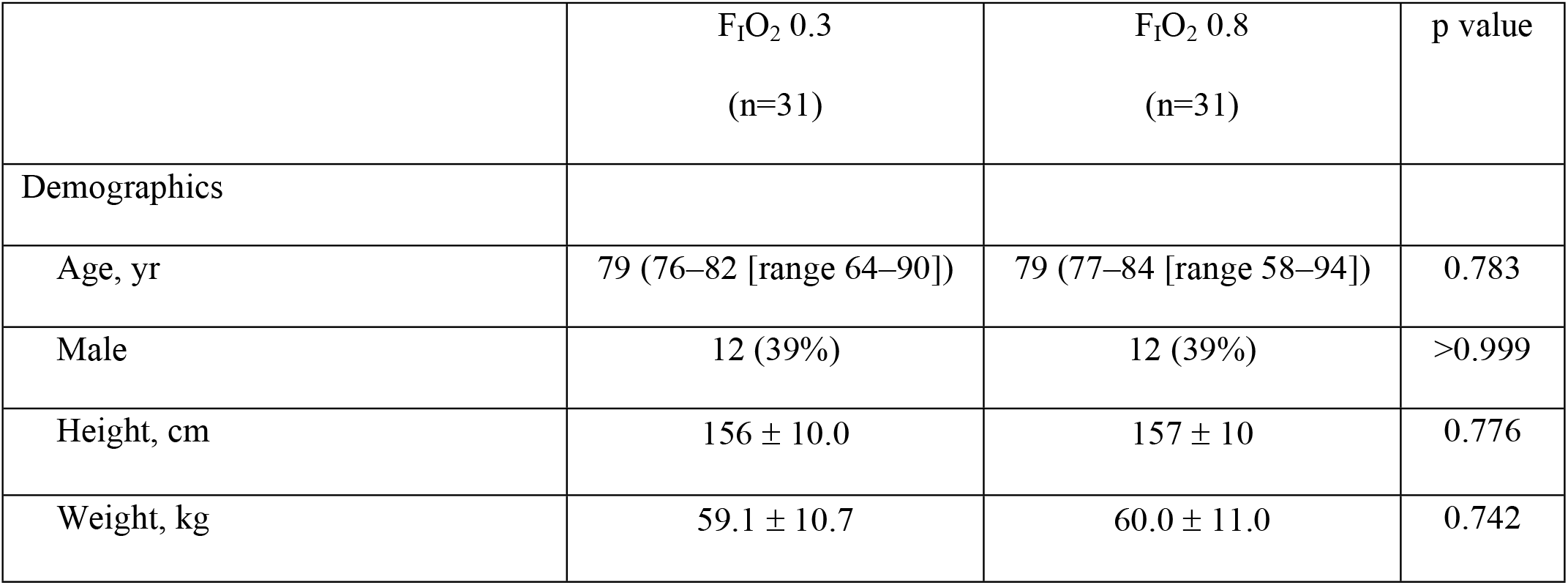
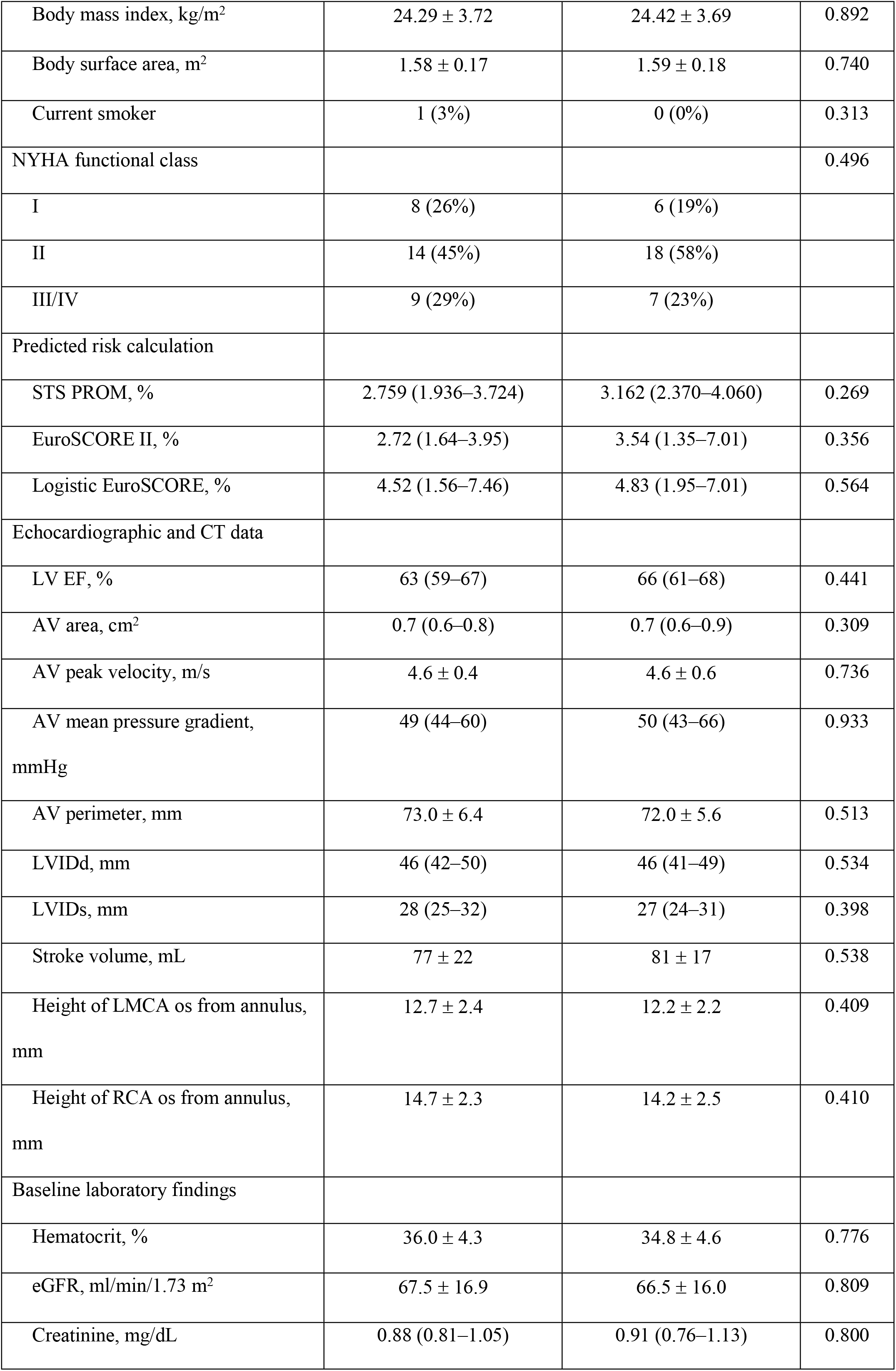
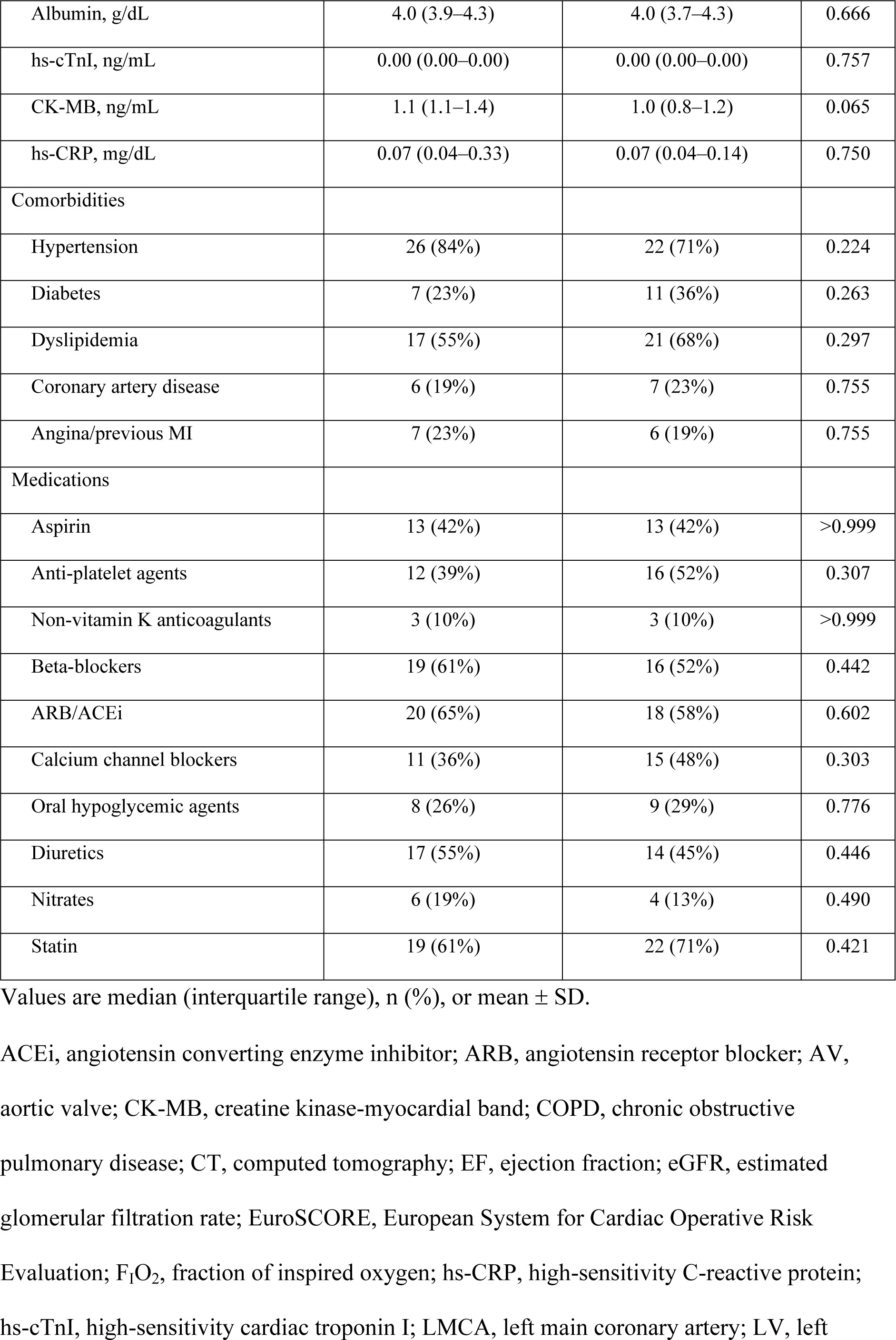
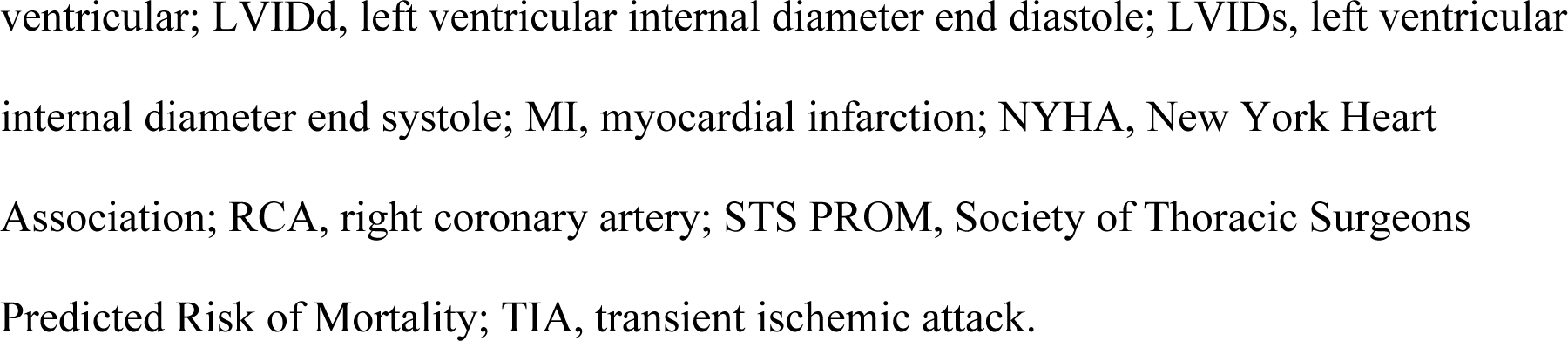
Baseline characteristics in patients undergoing transfemoral transcatheter aortic valve implantation.

**Table 2.** Valve characteristics and procedural variables in patients undergoing transfemoral transcatheter aortic valve implantation.

|  | F<sub>I</sub>O<sub>2</sub> 0.3<br>(n=31) | F<sub>I</sub>O<sub>2</sub> 0.8<br>(n=31) | p value |
| --- | --- | --- | --- |
| Prosthetic valve type |  |  | 0.647 |
| Sapien III | 13 (42%) | 14 (45%) |  |
| Evolut Pro or R | 15 (48%) | 12 (39%) |  |
| Lotus valve | 3 (10%) | 5 (16%) |  |
| Prosthetic valve size, mm |  |  | 0.698 |
| 23 | 6 (19%) | 9 (29%) |  |
| 25 | 1 (3%) | 2 (7%) |  |
| 26 | 18 (58%) | 14 (45%) |  |
| 29 | 6 (19%) | 6 (19%) |  |
| Paravalvular regurgitation |  |  | 0.112 |
| No | 12 (39%) | 13 (42%) |  |
| Trivial | 9 (29%) | 15 (48%) |  |
| Mild | 9 (29%) | 2 (7%) |  |
| Moderate | 1 (3%) | 1 (3%) |  |
| Procedure duration, min | 80 (70–95) | 80 (70–95) | 0.860 |
| Anesthesia duration, min | 140 (120–155) | 135 (120–160) | 0.888 |
| Amount of contrast media, mL | 250 (200–288) | 250 (210–326) | 0.404 |
| Transfusion of pRBC, unit | 0 (0–1) | 0 (0–1) | 0.675 |

Values are n (%) or median (interquartile range).
F<sub>I</sub>O<sub>2</sub>, fraction of inspired oxygen; pRBC, packed red blood cell.

During the procedure, PaO_2_, arterial oxygen saturation (SaO_2_), and SpO_2_ were higher in the F_I_O_2_ 0.8 than F_I_O_2_ 0.3 group (Fig 2). Two patients in the F_I_O_2_ 0.3 group required adjustment of F_I_O_2_ to 0.4 during the procedure because transient SpO_2_ <93% was observed. The mean left and right cerebral oximetry values were higher in the F_I_O_2_ 0.8 than F_I_O_2_ 0.3 group (Fig 2). For serial measurements, interactions between measurement time and group were significant for PaO_2_, SaO_2_, and the mean ScrbO_2_ (p<0.001, <0.001, and 0.032, respectively), but not for SpO_2_ (p=0.330).

**Fig 2.**
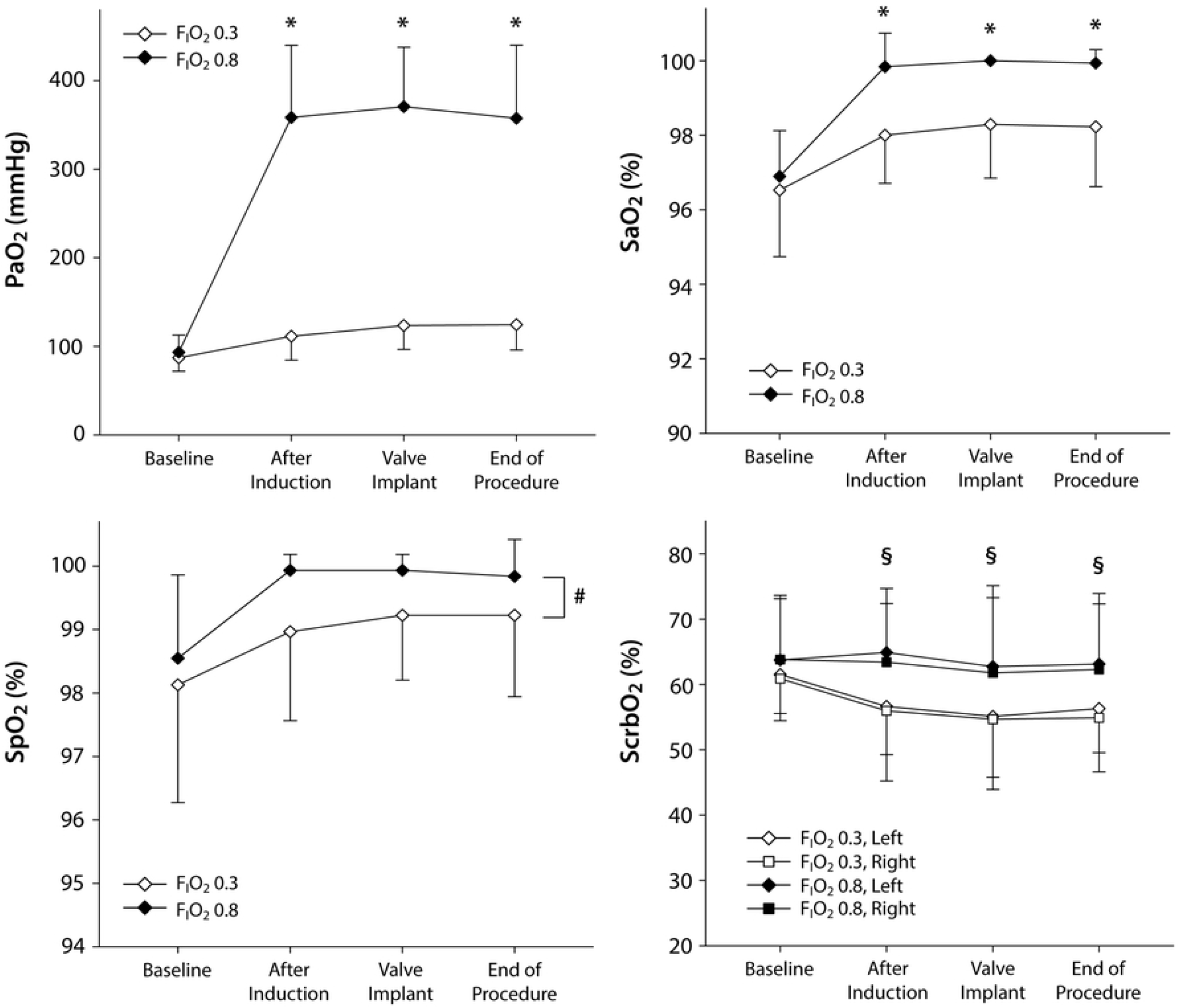
Changes in arterial oxygenation, pulse oxygen saturation, and cerebral oximetry in patients receiving a fraction of inspired oxygen of 0.3 or 0.8 during transcatheter aortic valve implantation. F_I_O_2_, fraction of inspired oxygen; PaO_2_, arterial partial pressure of oxygen; SaO_2_, arterial oxygen saturation; SpO_2_, pulse oxygen saturation; ScrbO_2_, cerebral oxygen saturation. * p < 0.001 compared to the F_I_O_2_ 0.3 group. # p = 0.003 between the groups (mixed model). § p < 0.05 compared to the F_I_O_2_ 0.3 group.

The primary outcome, the AUC for serum hs-cTnI in the first 72 h post-TAVI, was higher in the F_I_O_2_ 0.8 than F_I_O_2_ 0.3 group (71.96 [35.38–116.34] vs. 42.66 [24.82–65.44] h·ng/mL), but the difference was not statistically significant (p=0.066) (Fig 3). The secondary outcome (AUC for CK-MB in the first 72 h post-TAVI) was also higher in the F_I_O_2_ 0.8 group, but not significantly (342.2 [195.4–485.2] vs. 257.6 [155.6–322.0] h·ng/mL; p=0.132) (Fig 3). The peak hs-cTnI and CK-MB levels during the first 72 h post-TAVI were also non-significantly higher in the F_I_O_2_ 0.8 group (1.79 [1.09–3.77] vs. 1.30 [1.00–1.58] ng/mL and 10.9 [5.7–15.6] vs. 7.5 [6.0–11.9] ng/mL; p=0.051 and 0.159, respectively). For periprocedural serial measurements, the group differences in hs-cTnI and CK-MB did not reach statistical significance (p=0.125 and 0.084, respectively; mixed model). The interaction between measurement time and group was not significant (p=0.200 and 0.096 for hs-cTnI and CK-MB, respectively). When including the five patients who completed the study protocol and were excluded from the final analysis due to elevated baseline hs-cTnI after randomization, there were no significant differences in the AUCs for hs-cTnI and CK-MB in the first 72 h post-TAVI between the groups (S1 Fig). After adjustment for various risk factors, there was no significant association between the F_I_O_2_ and AUC for hs-cTnI post-TAVI (OR: 3.644, 95% CI: 0.864–15.371; p=0.078).

**Fig 3.**
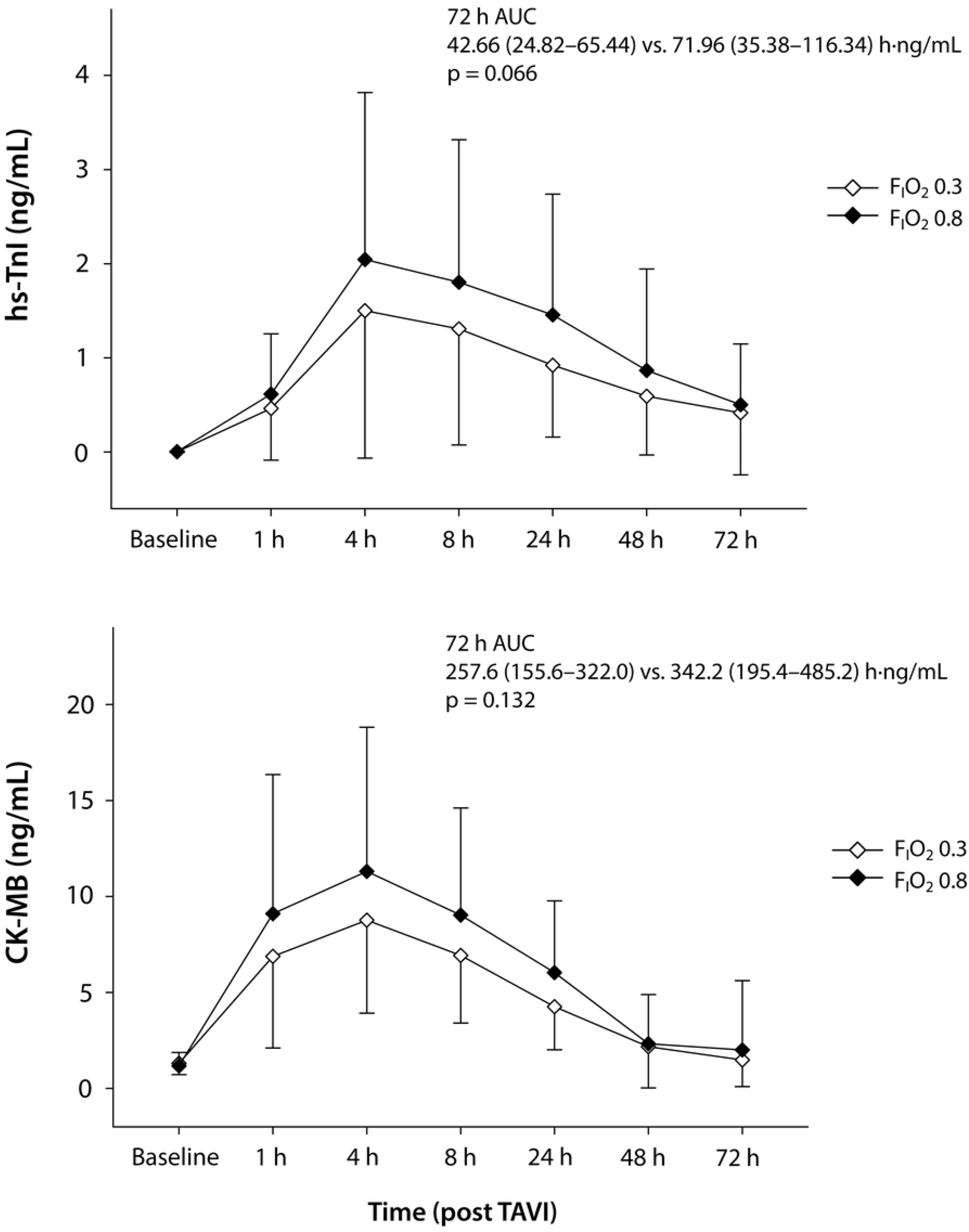
Changes in cardiac biomarkers in the first 72 h in patients who received a fraction of inspired oxygen of 0.3 or 0.8 during transcatheter aortic valve implantation. AUC, area under the curve; CK-MB, creatine kinase-myocardial band; hs-cTnI, high-sensitivity cardiac troponin I; TAVI, transcatheter aortic valve implantation.

The postprocedural incidence of AKI did not differ between F_I_O_2_ 0.3 and 0.8 groups (36% vs. 42%; p=0.602) (Table 3). However, AKR was more frequent in the F_I_O_2_ 0.3 than F_I_O_2_ 0.8 group (65% vs. 39%; p=0.042). Other postprocedural clinical outcomes, such as cardiovascular mortality or length of hospitalization, were comparable between the two groups (Table 3).

**Table 3.**
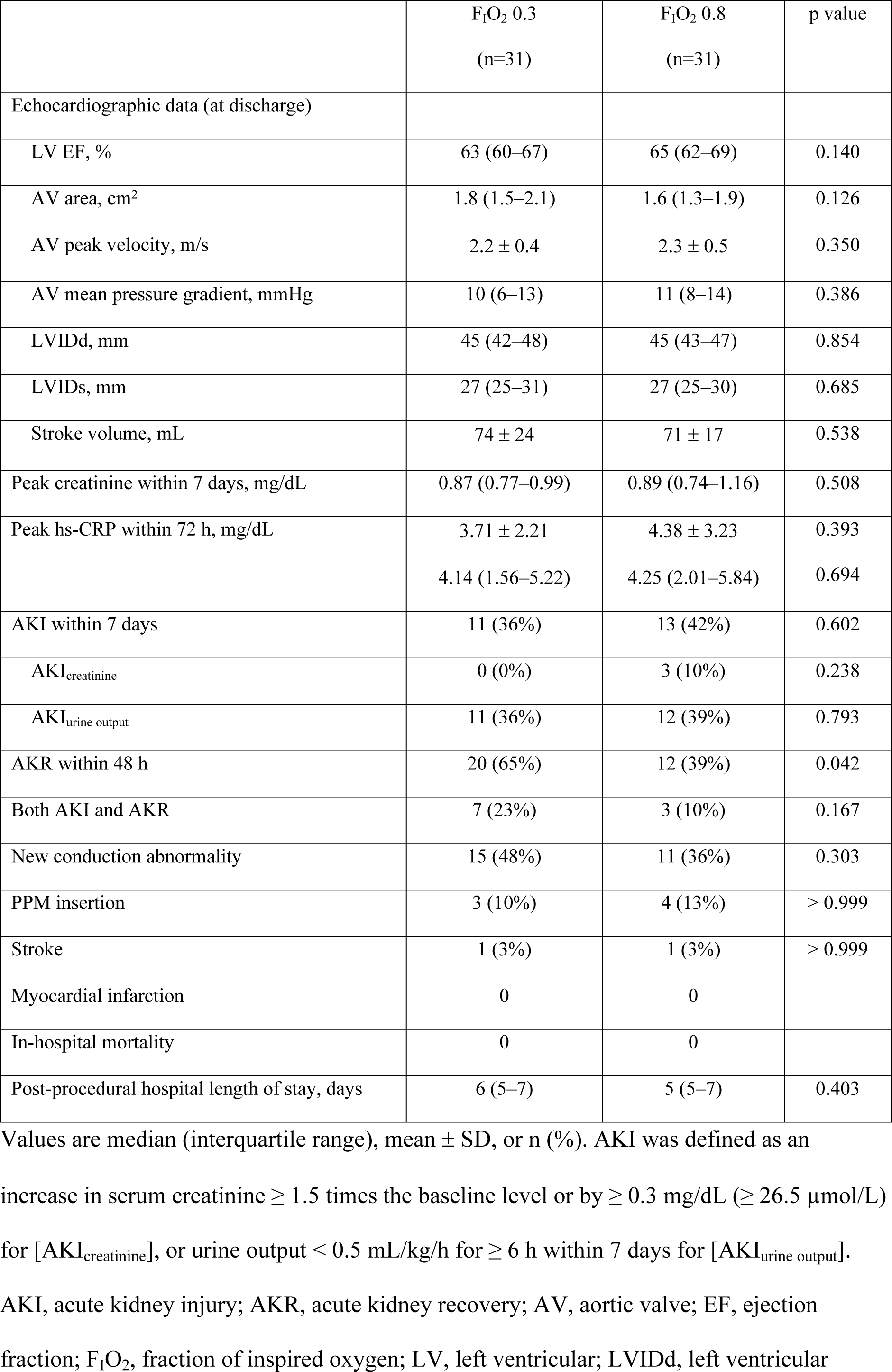
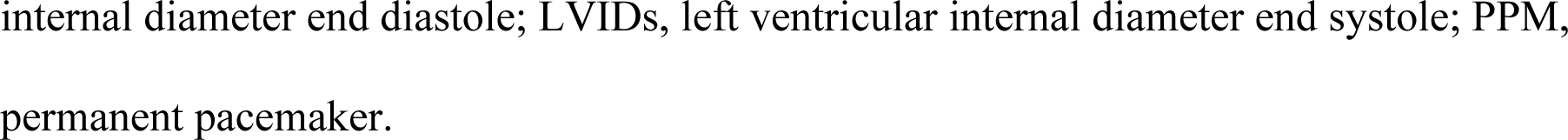
Postprocedural variables in patients received fraction of inspired oxygen 0.3 or 0.8 during transfemoral transcatheter aortic valve implantation.

## Discussion

Compared to the F_I_O_2_ 0.3 group, the F_I_O_2_ 0.8 group showed a greater postprocedural elevation of cardiac biomarkers, albeit without statistical significance. Postprocedural AKR was more frequent in the F_I_O_2_ 0.3 group. There was no difference in other periprocedural outcomes between the groups.

### Myocardial injury after TAVI

Even after successful TAVI, the cardiac biomarkers cTnI and CK-MB showed post-procedure increases despite prompt relief of transvalvular pressure overload, and periprocedural myocardial injury occurred along with transient deterioration in myocardial function [13]. Significant deterioration in the myocardial performance index, which implies both systolic and diastolic dysfunction, was observed immediately following TAVI [13]. New myocardial late enhancement with an ischemic pattern, indicating myocardial damage, was detected on cardiac magnetic resonance images following both balloon-expandable and self-expandable valve implantation [14].

Transient LV dysfunction and injury following TAVI seems to be partly influenced by procedural aspects of transcatheter valve deployment [15]. Procedure-related mechanical trauma during TAVI, including during balloon valvuloplasty, valve positioning, and prosthesis delivery, also plays a substantial role in myocardial damage [16]. Rapid ventricular pacing is used to temporarily reduce the LV output during pre-implantation balloon valvuloplasty and balloon-expandable valve implantation, and for post-implantation ballooning to reduce paravalvular leakage. Rapid ventricular pacing transiently reduces microvascular tissue perfusion and the flow index in small- and medium-sized vessels, and induces partial microcirculatory arrest and delayed recovery of microflow [17]. Subsequently, ventricular stunning and subsequent dysfunction may occur [15].

### Hyperoxia and myocardial injury

The role of oxidative stress in reperfusion injury is relatively well established. Abrupt oxidative reactions following reperfusion produce reactive oxygen species (ROS) from cardiomyocytes and endothelial cells, which amplifies local inflammatory responses and leads to a vicious cycle of ROS production [18]. The biological mechanism underlying the adverse effects of hyperoxia is related to the generation of ROS, specifically the superoxide anion, which has a negative impact on coronary blood flow and LV distensibility [19].

Hyperoxia can exacerbate oxidative stress and thereby worsen coronary vasoconstriction and myocardial injury. Interestingly, hyperoxic reperfusion limited myocardial necrosis in rodents with cardiovascular risk factors more so than in a normoxemic reperfusion group, while the reverse occurred in healthy rodents [18]. Similarly, in a preliminary canine MI model, administering 100% oxygen had beneficial effects on the myocardium by reducing myocardial infarct size and improving the EF after reperfusion compared to room-air ventilation [20].

However, in the AVOID trial, patients presenting with acute MI were randomized to receive oxygen 8 L/min via face mask or ambient air, and oxygen treatment increased myocardial injury and infarct size in patients without hypoxia [21]. In patients admitted to the intensive care unit (ICU), conservative use of oxygen, which aimed to maintain arterial oxygen tension within the physiological range, reduced ICU mortality compared to conventional use of oxygen [22]. In the large randomized DETO2X-AMI study, there was no difference in 1-year mortality or the peak cardiac troponin level between patients with suspected MI who received supplemental oxygen versus ambient air [23]. During general anesthesia for major non-cardiac surgery, there was no difference in myocardial injury— assessed using the AUC for high-sensitive troponin in the first 3 postoperative days— between F_I_O_2_ 0.3 and 0.8 administered intraoperatively and for 2 h after surgery [3]. In the ICU-ROX study, conservative use of F_I_O_2_ (≤0.21) during mechanical ventilation in adult ICU patients resulted in no difference in the number of ventilator-free days compared to standard administration of F_I_O_2_ [24]. In a more recent nationwide registry trial, high oxygen supplementation (6–8 L/min by face mask) resulted in no significant difference in the 30-day or 1-year mortality rate in patients with suspected acute coronary syndrome compared to low oxygen treatment [25].

### Myocardial injury and clinical outcomes after TAVI

Cardiac biomarkers elevation following surgery or intervention result from perioperative hemodynamic stress, inflammation, or oxygen supply and demand imbalance [26]. Periprocedural myocardial injury following TAVI was associated with a significantly increased risk of poor short- and long-term clinical outcomes, including 30-day and 1-year mortality, neurological events, and postprocedural permanent pacemaker implantation [27].

In our study, high oxygen tension during TAVI tended to increase the release of cardiac biomarkers in the first 72 h post-TAVI compared to the low oxygen tension group, but the difference was not significant. There was no clinical impact of the level of intraoperative oxygen tension during TAVI, except that recovery of kidney function was more common in the low-compared to the high-F_I_O_2_ group.

### Acute kidney recovery

Acute kidney recovery, which is a relatively recently described phenomenon, has been observed more frequently than AKI after both TAVI and surgical aortic valve replacement (SAVR) [28]. Following TAVI or SAVR, normalization of the aortic valve area, prompt relief of the trans-aortic pressure gradient, and normalization of post-stenotic flow abnormality occur. Regarding renal blood flow, a rapid increase in cardiac output and reduced LV afterload may cause abrupt hemodynamic changes in the early postprocedural period, such as renal congestion. In a recent prospective registry analysis, both AKI and AKR early after TAVI were independent predictors of cardiovascular mortality [29]. In our study cohort, 10 of the 62 (16%) patients met the criteria for both AKI and AKR during the study period (Table 3). Rapid changes in renal hemodynamics could have occurred in these groups, and both post-TAVI AKI and AKR may reflect a cardiorenal aspect of the extra-cardiac damage characterizing severe AS. Further studies are required to assess the relationship between renal circulatory changes and clinical outcomes in AS patients following TAVI.

### Study limitations

This study has some limitations. First, it was a single-center trial with relatively few patients, and was only powered for one surrogate cardiac biomarker, hs-cTnI. Although we observed a trend toward reduced hs-cTnI release and better postprocedural kidney recovery, we cannot definitely conclude that arterial oxygenation is beneficial for patients undergoing TAVI in terms of periprocedural myocardial and renal protection. Second, we included patients undergoing general anesthesia for transfemoral TAVI in this study. However, many patients undergo TAVI under conscious sedation or even local anesthesia, unless they are at very high periprocedural risk due to severely compromised cardiopulmonary function or the inability to maintain a stable supine position, for example. Nevertheless, the included patients had comparable characteristics regarding potential risk factors for postprocedural myocardial injury between the two groups. Future investigators could compare the oxygenation strategies of minimal supplemental oxygen and no supplemental oxygen, as in the treatment of acute MI patients without hypoxia, in terms of the likelihood of avoiding unnecessary periprocedural oxidative stress and protecting multiple organ systems, in patients undergoing TAVI under conscious sedation or local anesthesia. Third, we included relatively low-risk patients; we excluded those who were already hypoxemic or required supplemental oxygen, and those with acute coronary syndrome or renal failure. In high-risk patients with severe LV dysfunction or poor oxygenation, however, different supplemental oxygen strategies may have a differential impact on myocardial injury and other clinical outcomes. Therefore, further studies are required of high-risk patients, who may be more suitable candidates for TAVI. Lastly, there is lack of control subjects undergoing SAVR in evaluating oxygenation and perioprocedural myocardial injury in this study. As it is beyond the primary aim of the present study, future studies can be conducted regarding periprocedural oxygen content and myocardial injury in patients undergoing TAVI vs. SAVR.

## Conclusions

In conclusion, the F_I_O_2_ level did not have a significant effect on periprocedural myocardial injury following TAVI with general anesthesia. However, considering the marginal results, a benefit of low F_I_O_2_ during TAVI could not be ruled out.

## Data Availability

All relevant data are within the manuscript and its Supporting Information files.

## Acknowledgements

The authors gratefully acknowledge Sun-Young Jung, a statistician, who is collaborating with our institution, for her assistance in the statistical analysis and invaluable advice and comments.

## Supporting information

**S1 Fig. Changes in cardiac biomarkers in the first 72 h in patients including five patients who were excluded from the main analysis due to pre-procedural elevation of cardiac biomarkers after randomization and received a fraction of inspired oxygen of 0.3 or 0.8 during transcatheter aortic valve implantation**. AUC, area under the curve; CK-MB, creatine kinase-myocardial band; hs-cTnI, high sensitivity cardiac troponin I; TAVI, transcatheter aortic valve implantation.

**S1 CONSORT Checklist**.

